# Improvement in estimates of GFR by using fat-free mass as compared to body weight in Indians: pilot study

**DOI:** 10.1101/2023.05.25.23289723

**Authors:** Anjali R Kulkarni, Chittranjan S Yajnik, Lavanya Sampathkumar, T R Dilip

## Abstract

**Background:** Estimated glomerular filtration rate (eGFR) may be calculated by different equations with variable accuracy. The accuracy of creatinine-based eGFR equations may vary across ethnic groups. These are also influenced by differences in body composition. Many populations have higher adiposity for a given body mass index, this disparity is very striking in Indians and has led to the description of a ‘thin-fat’ Indian.

We undertook a pilot study to compare eGFR estimated from clinical equations using fat-free mass instead of total body weight, in healthy Indians.

**Methods and results:** We studied 26 healthy Indian volunteers (11 men, 15 women), aged 49 (34,55) years [Median (Min, Max)]. We recorded vitals, anthropometric and biochemical measurements. eGFR values were estimated by Cockcroft-Gault (CG-BSA), Modification of Diet in Renal Disease (MDRD), and Chronic Kidney Disease Epidemiology Collaboration (CKD-EPI) formulae. eGFR values based on the CG-BSA equation were re-calculated after replacing coefficient of total body weight with fat-free mass and muscle mass (obtained by BIA) and lean mass (obtained by DEXA scan).

We used Tc 99m DTPA renogram for GFR measurement as the gold standard.

Median eGFR using conventional formulae overestimated the GFR when compared to mGFR (81 ml/min/1.73m^2^); CG-BSA equation (99 mL/min/1.73 m^2^); EPI-CKD equation (97 mL/min/1.73 m^2^) and MDRD equation (89 mL/min/1.73 m^2^).

Of the CG-BSA formulae, eGFR using total Body weight is the farthest from gold standard. And estimated GFR using fat free mass had closest median (67 mL/min/1.73 m^2^) to the gold standard (79 mL/min/1.73 m^2^)

Bland-Altman analysis showed the Mean difference of Fat free mass-based CG-BSA formula from the gold standard was the lowest of all four CG formulae (−12.009, CI -19.266 to -4.752); showing that it had the best agreement with the gold standard of the four CG-BSA formulae. Also, it showed that the mean difference of MDRD formula from gold standard is the lowest (9.191, CI 1.008 to 17.375) followed by Cockcroft Gault formula using Fat Free mass (−12.009, CI -19.266 to -4.752).

Mean difference of EPI formula (15.151, CI 8.182 to 22.120) and Cockcroft Gault formula using Muscle mass (−15.809, CI -22.756 to -8.861) follow, with very similar Limits of Agreement (LOA).

**Conclusion:** This pilot study showed that existing conventional eGFR equations CG-BSA, MDRD, and CKD-EPI overestimate eGFR in healthy Indian subjects. The conventional CG-BSA had the least agreement with measured GFR by the gold standard. However, the replacement of coefficient of total body weight by FFM, MM and LM in the CG-BSA formula improved the estimates of eGFR in healthy Indian volunteers.

**Significance Statement:** Estimation of glomerular filtration rate (eGFR) is an important practice in clinical medicine and there are various equations available to obtain it. eGFR varies with serum creatinine and body composition which is different for the Indian population as compared to the African and Caucasian population. There has been no validation of the commonly used equations in the Indian population. Usage of coefficients based on body composition such as fat-free mass, lean mass or muscle mass to calculate the eGFR may be explored in the Indian population in view of the presence of lower muscle mass and higher adiposity in them, as per the concept of “thin-fat Indian”

## Introduction

Assessment of kidney function is a key part of routine medical evaluation and plays a vital role in accurate diagnosis, appropriate dosing of medicines, and pre-operative assessment of the patients, amongst other uses. Glomerular filtration rate (GFR) is considered one of the best available indices of kidney function in health and disease. Since the measurement of GFR is complex and time-consuming, it is not measured routinely in clinical practice.

The normal GFR varies considerably with age, sex, body size, physical activity, diet, and pharmacologic therapy ^1^ and is usually estimated based on measurement of creatinine clearance. Such estimated glomerular filtration rates (eGFR) can be calculated by several equations; the most commonly used ones are Cockcroft-Gault (CG) equation, the Modification of Diet in Renal Disease (MDRD) Study equation, and the Chronic Kidney Disease Epidemiology Collaboration (CKD-EPI) equation^. 2^

The question of which is the best eGFR equation to assess GFR values remains unresolved in Indians, with wide variation in the usage of equations.

The factors which stand out in Indians as compared to African and Caucasian populations, are the differences in body composition and low muscle mass ^3^. A few studies have tried to evaluate these eGFR equations for their ability to predict kidney function, with respect to age, gender and dietary protein intake (factors known to affect serum creatinine values).

These studies have variable outcomes^4,5^. However, not many studies have validated the use of these equations in the Indian population.

The use of coefficients based on parameters of body composition such as Fat Free mass or muscle mass have often been postulated for the different eGFR equations although they have not been compared with the CG, MDRD and CKD-EPI equations. The aim of this pilot study is to compare the performance of conventional creatinine-based GFR equations with eGFR equations using Fat-Free mass/ lean mass as coefficients in healthy Indian subjects.

## Materials and Methods

### Participant subjects

We screened 500 apparently healthy volunteers who attended various health awareness programs. All the volunteers who were considered for inclusion in the study were informed about the study protocol. After excluding known patients of diabetes, hypertension, renal diseases and cardiovascular diseases, informed consents were obtained. Healthy pregnant women were also excluded from the study

150 volunteers who consented for participation, were subjected to thorough clinical and biochemical evaluation to rule out any associated medical illness.

Total 26 volunteers in the age group of 18 to 65 years who had no major medical illness, were enrolled in this pilot study for further evaluation.

### Measurements

The anthropometrical measures were made by two trained observers after 8 hours of overnight fasting.

Height was measured to the nearest 0.1 cm using a stadiometer (Detecto-medic, Detectoscales Inc, Brooklyn. N. Y.USA). Body weight was recorded to the nearest 0.1 kg using a portable electronics scale (Accuweigh, Model PFE, Mumbai).

#### Body composition analysis

was conducted using two methods

1. **DEXA scan:** Lean body mass (LBM) was measured by DEXA scan (Lunar prodigy, GE Healthcare) for all the participants.
2. **Bioelectrical Impedance Analysis** (BIA): Fat-free mass (FFM) and Muscle mass (MM) were measured by Bioelectrical Impedance analyzer (Tanita MC 780 Multi-Frequency Segmental Body Composition Monitor, Japan), for all the participants ^6^.

#### Isotope Renal Scan

Tc-99 m DTPA (diethylene-triamine-pentaacetate) renogram was done, in 25 subjects, in well-hydrated state to measure GFR. The tracer washout was measured at 30 minutes for both kidneys without any diuretic use and total GFR was measured in each subject. One female participant did not agree to undergo Isotope renal scan and was not considered for further statistical analysis. The mGFR by this method has been considered the gold standard against which all other methods have been compared.

#### Biochemical tests

Serum creatinine measurements were done using the Modified Jaffe method (Kinetic alkaline picrate method, Konelab 30 Prime – Fully automatic biochemistry analyzer. Thermo Scientific, USA). Plasma glucose, HbA1C, serum uric acid, BUN, and lipids were by standard laboratory methods to rule out any associated medical illness in the selected study population.

### eGFR calculation

Estimation of GFR was done in all the participants by using Cockcroft-Gault adjusted for body surface area (CG-BSA), ^7,8^ Modified diet in renal diseases (MDRD), Chronic Kidney Disease Epidemiology (CKD-EPI) equation (Table 1).

**Table 1.** eGFR Equations used in the study

|  |  |
| --- | --- |
| Cockcroft-Gault (CG-BSA) | $eGFR (CG) = \frac{[(140 - age) * weight * 0.85 \text{ if female}]}{72 * SCr}$<br>*adjusted for BSA<sup>#</sup> by (1.73 m<sup>2</sup>)/BSA* |
| Modified diet in renal diseases (MDRD) | $eGFR (MDRD) = (186 * (SCr)^{-1.154} * (age)^{-0.203} * (0.742 \text{ if female}))$ |
| Chronic Kidney Disease Epidemiology (CKD-EPI) | $eGFR (CKD - EPI) = \left( 141 * \min\left(\frac{SCr}{\kappa}, 1\right)^{\alpha} * \max\left(\frac{SCr}{\kappa}, 1\right)^{-1.209} * 0.993^{age} * (1.018 \text{ if female}) \right)$ |

#BSA= Body Surface area SCr= Serum Creatinine values

eGFR values based on the CG-BSA equation were re-calculated after replacing total body weight with fat-free mass and muscle mass (obtained by BIA) and lean mass (obtained by DEXA scan)

All the mGFR values were measured using DTPA Tc 99 Isotope renal scan in all participants except one female patient, who was unwilling to undergo the scan.

### Statement of Ethical Approval

The study was conducted in accordance with the ethical principles to be followed for medical research adopted in the Declaration of Helsinki and the Declaration of Istanbul and was approved by the Institutional Ethics Committee of the Bhabha Atomic Research Centre Hospital, Mumbai, India.

### Statistical Analysis

Median values including maximum and minimum values are computed and reported for the following attributes of the study participants: age, body mass, blood sugar, lipid profile, blood pressure, uric acid, and creatinine levels.

Estimates of GFR obtained by various conventional as well as modified equations and mGFR obtained by Isotope renal scan were compared using Bland-Altman analysis to understand correlation and agreement. Bias was expressed as the mean difference between m GFR and eGFR. The 95% limits of agreement (LOA) were calculated as mean bias ± 1.96 x standard deviation.

Data were analyzed using Stata 16 software (StataCorp 2019 Stata Statistical Software: Release 16. College Station, TX: StataCorp LLC).

## Results

We included a total of 26 healthy volunteers (11 males, 15 females) in the pilot study after applying strict inclusion and exclusion criteria and a thorough clinical examination, and biochemical evaluation. Table 2 shows the characteristics of the study population; the median age was 49 years and the median weight was 67 kilograms. The median creatinine value was 0.9 mg/dl, Median Fasting blood sugar was 94 mg/dl and Median HbA1C was 6%.

**Table 2.**
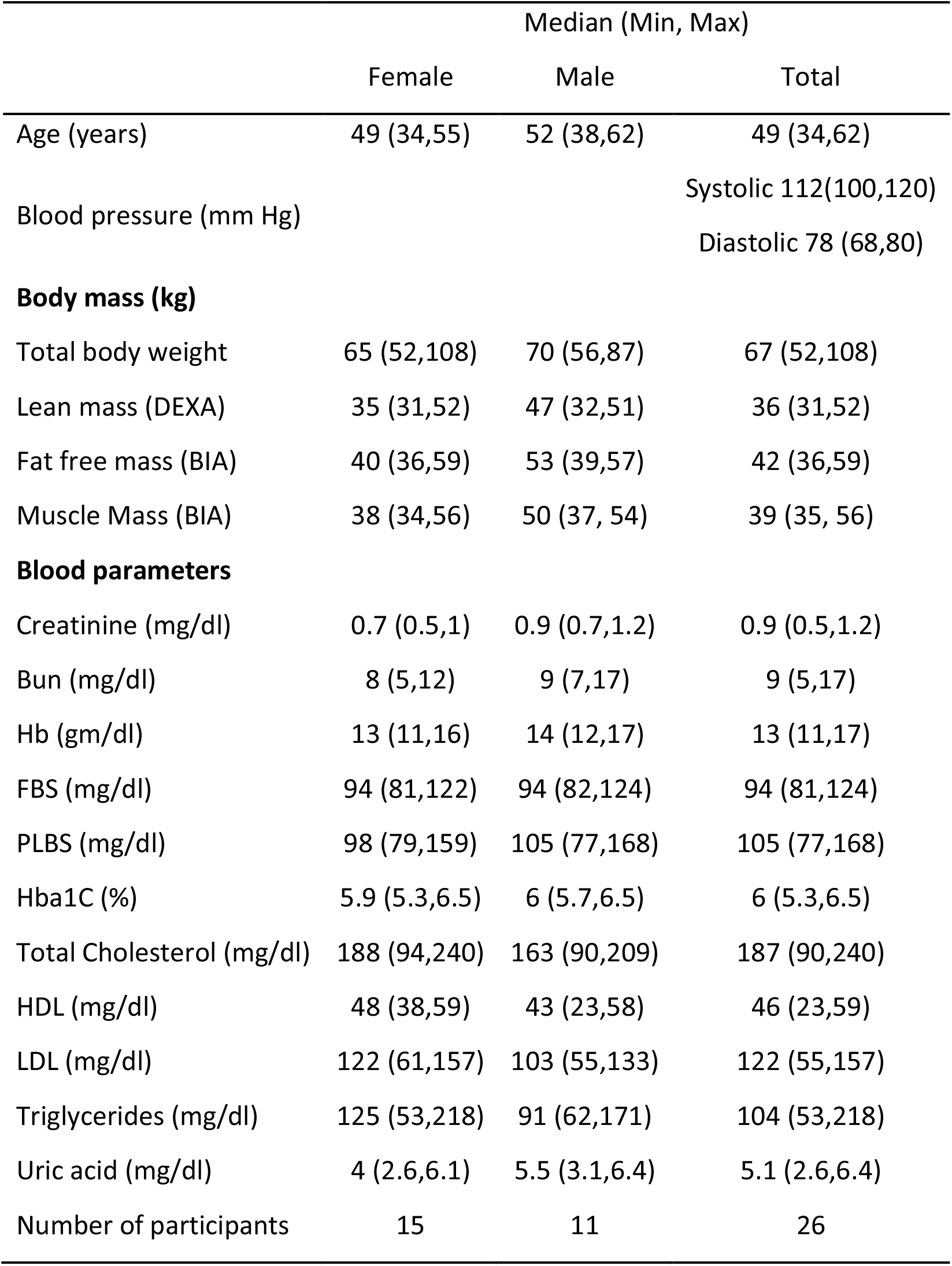
Characteristics of Study population

Table 3. (Estimated GFR) and mGFR (Measured GFR) in the study population.In our study, the median eGFR was highest when calculated using total body weight in the CG-BSA equation (99 ml/min/1.73 m^2^), followed by the one obtained by using CKD-EPI equation (97 ml/min/1.73 m^2^) and MDRD equation (89 ml/min/1.73 m^2^) respectively.

**Table 3.**
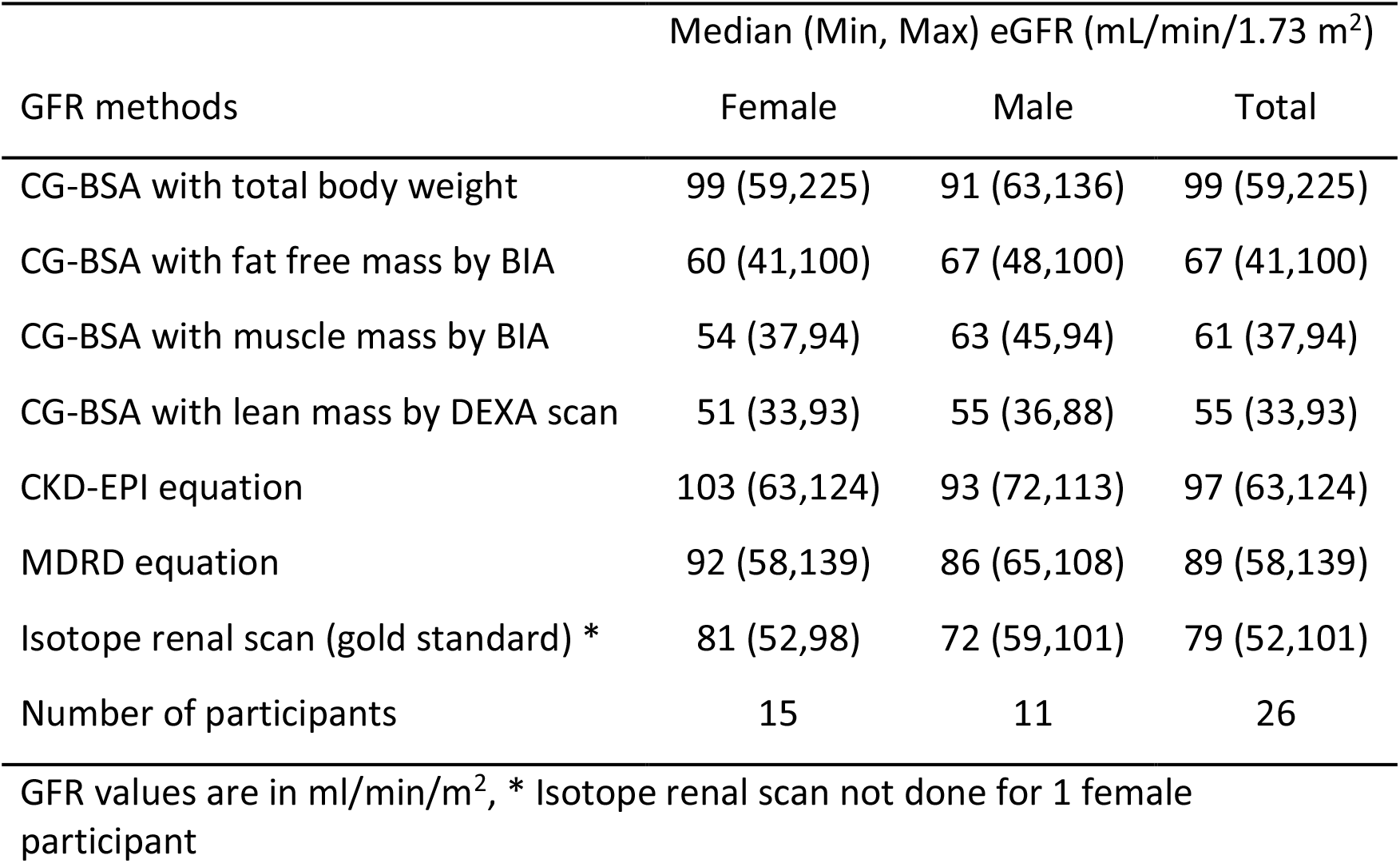
Estimates of GFR by equations under study

The lowest median value of eGFR was obtained in case of the equation using Lean Mass, followed by Muscle Mass and Fat-Free Mass respectively.

We observed that eGFR values by all three conventional equations were found to be higher as compared to measured GFR by Isotope renal scan. Thus, these existing creatinine-based equations significantly overestimated eGFR in our study as compared to measured GFR. Estimation of eGFR by CG-BSA using total Body weight is the farthest from gold standard.

The median value of eGFR was higher for females than males, if it was computed using total body weight, CKD-EPI equation, MDRD equation, and by DTPA Tc 99 scan. On the contrary, eGFR was observed to be higher for males than females if computation is based on coefficients of body composition like fat-free mass, muscle mass and lean mass.

Table 4 and Figure 1 give the Bland-Altman comparison, done for all 6 eGFR equations compared to Isotope Renal scan as the gold standard. The Mean difference of MDRD equation from gold standard is the lowest (9.191, CI 1.008 to 17.375) followed by Cockcroft Gault equation using Fat-Free mass (−12.009, CI -19.266 to -4.752).

**Table 4.** Bland-Altman comparison of difference in various measures of eGFR with gold standard (N=25)

|  | Mean difference (95 % CI) | Pitman's Test of difference in variance (r) | Significance level |
| --- | --- | --- | --- |
| CG-BSA with total body weight | 23.071 (CI 10.062 to 36.080) | 0.806 | 0 |
| CG-BSA with fat free mass | -12.009 (CI -19.266 to -4.752) | 0.354 | 0.082 |
| CG-BSA with muscle mass | -15.809 (CI -22.756 to -8.861) | 0.300 | 0.146 |
| CG-BSA with lean mass | -21.729 (CI -28.715 to -14.742) | 0.310 | 0.132 |
| CKD - EPI equation | 15.151 (CI 8.182 to 22.120) | 0.368 | 0.071 |
| MDRD equation | 9.191 (CI 1.008 to 17.375) | 0.477 | 0.016 |

**Figure 1.**
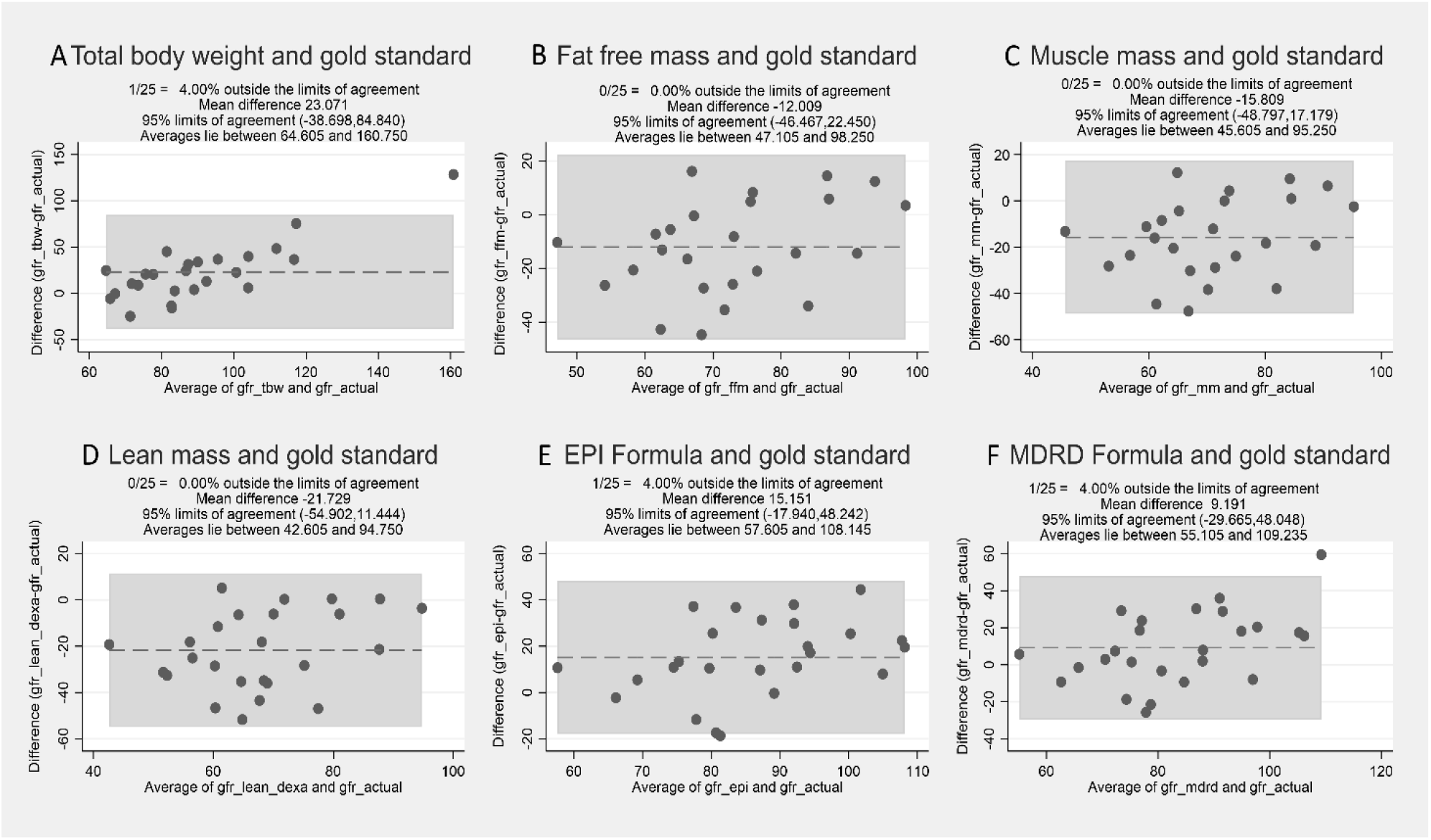
Bland-Altman comparison of difference in various measures of eGFR with gold standard (N=25) Mean differences of all 6 eGFR equations in the study have been plotted against gold standard mGFR. CG-BSA with total body weight (Panel A) had the largest mean difference (23.071 ml/min/1.73m^2^) and MDRD (Panel F) has the smallest mean difference (9.191 ml/min/1.73m^2^). Of the 4 CG-BSA equations (Panels A – D), CG-BSA with FFM has the lowest mean difference and narrowest Limits of agreement. (−12.009 ml/min/1.73m^2^ (CI -19.266 to -4.752).

The mean difference of CKD-EPI equation (15.151, CI 8.182 to 22.120) and Cockcroft Gault equation using Muscle mass (−15.809, CI -22.756 to -8.861) follow, with very similar Limits of Agreement (LOA).

Of the four Cockcroft Gault (CG-BSA) equations, the value of eGFR using fat-free mass had the closest median (67 ml/min/1.73 m^2^) to the gold standard (79 ml/min/1.73 m^2^). Also, the Mean difference of the Fat-free mass CG-BSA equation from the gold standard (−12.009, CI - 19.266 to -4.752) was the lowest of all four CG-BSA equations, showing that it has the best agreement with the gold standard of the four CG-BSA equations.

In our study, the overall estimation of eGFR by MDRD is closest to the gold standard when Median value is compared. But the Limits of agreement for eGFR by CKD-EPI equation is narrower. This showed a conflicting outcome.

## Discussion

This pilot study was conducted on healthy Indian volunteers for evaluation of eGFR using various coefficients of body composition, in view of ethnic metabolic differences.

The study showed that the existing creatinine-based eGFR equations, significantly overestimate GFR when compared with mGFR by DTPA Tc 99 renal scan.

CG-BSA equation agreement analysis (Table 4), with isotope renal scan measurement as the gold standard, indicates that the conventional and most commonly used CG-BSA equation using total body weight.

While considering values of median eGFR and limits of agreement for MDRD and CKD-EPI equations, we could not get conclusive results. Further studies are required for the same.

As per the literature, there have been constant efforts to modify the conventional CG equation for the relevant population.

Rolin and co-workers in 1984, concluded that the original Cockcroft-Gault equation was an inaccurate predictor of GFR and correction of the equation for patient physical parameters improved its accuracy for GFR prediction. ^9^

The study by Vivek Kumar et.al. showed that the existing creatinine-based GFR estimating equations significantly overestimate GFR in the Indian population, which could be linked to the low muscle mass and vegetarian diet. ^10^

Prashanth Rajagopalan et al in their study attempted to establish a normal range of serum creatinine and cystatin C values for the Indian population and showed that in young healthy Indian adults, eGFR and kidney volume vary by BMI and sex.^11^

As the influence of muscle mass on serum creatinine levels is well-known, some studies showed that muscle mass affects serum and urinary creatinine, thus affecting the value of eGFR. ^12–14^

The Cockcroft-Gault equation is an estimate of creatinine clearance originally developed in a population of 236 Canadian patients, 209 of whom were male, however, in 1992, Gault and co-workers suggested the use of ideal body weight. ^15^ The results of a study on adult patients with chronic renal disease suggested that although body weight is already included in the Cockcroft-Gault equation, correction for body surface area further improves the accuracy of these equations ^7^.

Indian population is known to have higher adiposity for a given body mass index, this disparity has led to the description of a ‘thin fat Indian’. ^16^ They also have less proportion of muscle mass and more adiposity for a given BMI. ^17^

Though these equations are most commonly used in clinical practice, they are not validated in the Indian population. Indians may have lower serum creatinine levels and physiologically lower eGFR values in view of low muscle mass. ^18^ It has been shown that Indians may have low basal GFR as compared to the Western population.^19^ As per the first report of the Indian CKD registry, the emergence of diabetic nephropathy is an important cause of kidney disease. In addition to this, a significant proportion of patients have CKD of unknown etiology at a younger age. ^20^

The current pilot study on a comparatively younger mean age group (median age: 49 years) emphasizes the need for a ‘new normal of GFR range” for the Indian population as Cardio-metabolic risk is high in Indians manifesting at an early age. ^21^

Strengths of the current pilot study on healthy volunteers include that it took into account the ‘Thin-Fat-Indian” phenotype for estimation of GFR values considering the ethnic differences in body composition. We have used body composition parameters like FFM, LM, and MM for eGFR evaluation which are validated with Gold standards like isotope renal scan and DEXA scan. We have also utilized Bioelectrical Impedance Analysis (BIA) which is a relatively simple, inexpensive, and non-invasive bedside technique. To the best of our knowledge and literature survey, we could not find a similar study for eGFR equations using coefficients of body composition and the use of Bio-electrical Impedance Analysis (BIA) with comparable results like DEXA in Indian clinical practice.

In conclusion, the current pilot study showed that the existing conventional eGFR equations CG-BSA, MDRD and CKD-EPI overestimated eGFR in healthy Indian subjects. The conventional CG-BSA had the least agreement with measured GFR by the gold standard. The replacement of the coefficient of total body weight by FFM, MM, and LM in the CG-BSA equation improved the estimates of eGFR in healthy Indian volunteers. It is important to have large-scale studies to validate the conventional creatinine-based equations in the Indian setting.

We believe that this pilot study will reinvigorate the concept of using FFM/ LM in clinical practice for Indian patients and validate the existing eGFR equations.

## Limitations of the study

This study is conducted as a pilot project on 26 healthy Indian Volunteers as it required interventional procedures like Isotope Renal Scan and DEXA in normal persons.

It needs to be ascertained if these modified CG-BSA equations with coefficients of body composition perform equally well in patients with chronic kidney disease and other kidney conditions and larger studies will be necessary to fully validate the use of body composition measurements in clinical practice to calculate eGFR.

## Data Availability

All data produced in the present study are available upon reasonable request to the authors
All data produced in the present work are contained in the manuscript

## Author Contributions

### Conception and Design of study

Anjali R Kulkarni, Chittranjan S Yajnik,

### Acquisition of data

Anjali R Kulkarni, Lavanya Sampathkumar

### Analysis and Interpretation of data

Anjali R Kulkarni, Lavanya Sampathkumar, Dilip T R

### Drafting the manuscript

Anjali R Kulkarni, Chittranjan S Yajnik, Lavanya Sampathkumar

### Revising the manuscript critically for important intellectual content

Anjali R Kulkarni, Chittranjan S Yajnik, Lavanya Sampathkumar, Dilip T R

### Approval of the version of the manuscript to be published

Anjali R Kulkarni, Chittranjan S Yajnik, Lavanya Sampathkumar, Dilip T R

## Acknowledgments

Dr. Deepak Patkar

(Director, Medical Services, Head, Department of Radiology, Nanavati Super Speciality Hospital, Mumbai) for extending services and support for DEXA and Isotope renal scans.

## Disclosures

None

## Funding

None

## Notes

### Competing Interest Statement

The authors have declared no competing interest.

### Funding Statement

This study did not receive any funding

### Author Declarations

The study was approved by the Institutional Ethics Committee of the Bhabha Atomic Research Centre Hospital, Mumbai, India.

